# Examining the Association between the Gastrointestinal Microbiota and Gulf War Illness: A Prospective Cohort Study

**DOI:** 10.1101/2022.02.24.22270180

**Authors:** Ashley Kates, Julie Keating, Kelsey Baubie, Nathan Putman-Buehler, Lauren Watson, Jared Godfrey, Courtney L. Deblois, Garret Suen, Dane B. Cook, David Rabago, Ronald Gangnon, Nasia Safdar

## Abstract

Gulf War Illness (GWI) affects 25-35% of the 1991 Gulf War Veteran population. Patients with GWI experience pain, fatigue, cognitive impairments, gastrointestinal dysfunction, skin disorders, and respiratory issues. In longitudinal studies, many patients with GWI have shown little to no improvement in symptoms since diagnosis. The gut microbiome and diet play an important role in human health and disease, and preliminary studies suggest it may play a role in GWI. To examine the relationship between the gut microbiota, diet, and GWI, we conducted an eight-week prospective cohort study collecting stool samples, medications, health history, and dietary data. Sixty-nine participants were enrolled into the study, 36 of which met the case definition for GWI. The gut microbiota of participants, determined by 16S rRNA sequencing of stool samples, was stable over the duration of the study and showed no within person (alpha diversity) differences. Between group analyses (beta diversity) identified statistically significant different between those with and without GWI. Several taxonomic lineages were identified as differentially abundant between those with and without GWI (n=9) including a greater abundance of Lachnospiraceae and Ruminococcaceae in those without GWI. Additionally, there were taxonomic differences between those with high and low HEI scores including a greater abundance of Ruminococcaceae in those with higher HEI scores. This longitudinal cohort study of GWVs found that participants with GWI had significantly different microbiomes from those without GWI. Further studies are needed to determine the role these differences may play in the development and treatment of GWI.

## Introduction

Gulf War Illness (GWI) is a devastating chronic multi-symptom syndrome impacting 25-35% of the 700,000 coalition troops deployed during the 1990-91 Gulf War.(1),(2) Following their return from the Persian Gulf, Veterans experienced a unique pattern of symptoms across multiple physiologic domains with no identifiable cause. Over the last 30 years, a number of theories as to the cause of GWI have arisen with consensus being that troops were exposed to environmental contaminants – most likely pyridostigmine bromide (PB) or chemical nerve agents – during their deployment during Operations Desert Shield and Desert Storm.(3) Individuals with GWI experience musculoskeletal pain, fatigue, neurological symptoms (e.g., memory and cognitive issues, difficulty sleeping, depression, dizziness, headaches), rashes and other skin disorders, respiratory symptoms (e.g., persistent cough and difficulty breathing), as well as gastrointestinal issues (e.g., diarrhea, nausea, vomiting, and Irritable Bowel Syndrome).(4) Few effective treatments have been found, and patients have experienced little to no improvement in their symptoms over time.(5) Additionally, these patients are aging faster and developing additional comorbidities such as heart disease, diabetes, and arthritis at younger ages than the general population.(6)

While several case definitions for GWI exist, the Kansas case definition is most frequently reported; it is also one of the recommended case definitions by the Institute of Medicine.(4),(7) According to the Kansas case definition, patients must have moderately severe symptoms in three or more of the six symptom domains (chronic fatigue, neurological symptoms, joint or muscle pain, gastrointestinal disturbance, respiratory symptoms, and skin problems) for at least 6 months that are not explained by any other medical or psychiatric condition. Using these criteria, roughly one-third of GW Veterans (GWVs) meet the definition of GWI.(4)

Research to date primarily focuses on neurologic symptoms of GWI; however, gut dysfunction is also a commonly reported symptom of GWI patients.(8) Veterans with GWI related gut dysfunction experience pain, nausea, vomiting, bloating, and diarrhea similar to Irritable Bowel Syndrome (IBS) and Myalgic Encephalomyelitis/Chronic Fatigue Syndrome (ME/CFS). Disturbances to the gut microbiota may play a role in disease pathogenesis of both ME/CFS and IBS.(9),(10) Given the similarities between these syndromes and GWI, researchers have begun to consider the role of the gut microbiome role in GWI; preliminary studies show significantly different gut microbiomes between healthy controls and those with GWI in both animals and humans. Mice treated with PB and corticosterone had over 100 different operational taxonomic units (OTUs) than controls (p=0.005).(11) Similarly, mice treated with PB and permethrin had reduced alpha diversity (p <0.001) compared to controls; high fat diets further reduced alpha diversity while returns to normal feed provided partial recovery, indicating the potential role of diet in GWI symptom exacerbation.(12) A small pilot study of GWVs with GWI found those with GWI and gut symptoms had significantly different microbiomes compared to both those with GWI and no gut symptoms and healthy controls including higher abundances of Bacteroidetes, Actinobacteria, Erysipelotrichaceae, and Bifidobacteriaceae.(13)

We undertook a prospective cohort study to assess the relationship between GWI and the gut microbiome in GWVs with GWI and healthy controls. We hypothesized those with GWI would have less diverse microbiomes and higher Bacteroidetes abundance than healthy controls.

## Methods

We conducted an eight-week prospective cohort study assessing the gut microbiomes of GWVs. A detailed description of the study methodology has previously been published.***(14)***

### Study population, recruitment, and consent

Veterans meeting deployment criteria were identified from VA databases and were recruited via mailed invitation letters. Following invitation letters, GWVs interested in participating completed a phone screening to determine eligibility. Inclusion and exclusion criteria can be found in Table 1. Written informed consent was obtained at the enrollment visit by a trained team member prior to initiating additional study procedures. Participants received $50 for completing the enrollment visit and an additional $50 for the close-out visit. This study and all associated protocols were approved by the University of Wisconsin Madison Health Sciences Institutional Review Board (ID No. 2017-1212) and the Madison VA Research and Development Committee.

**Table 1.**
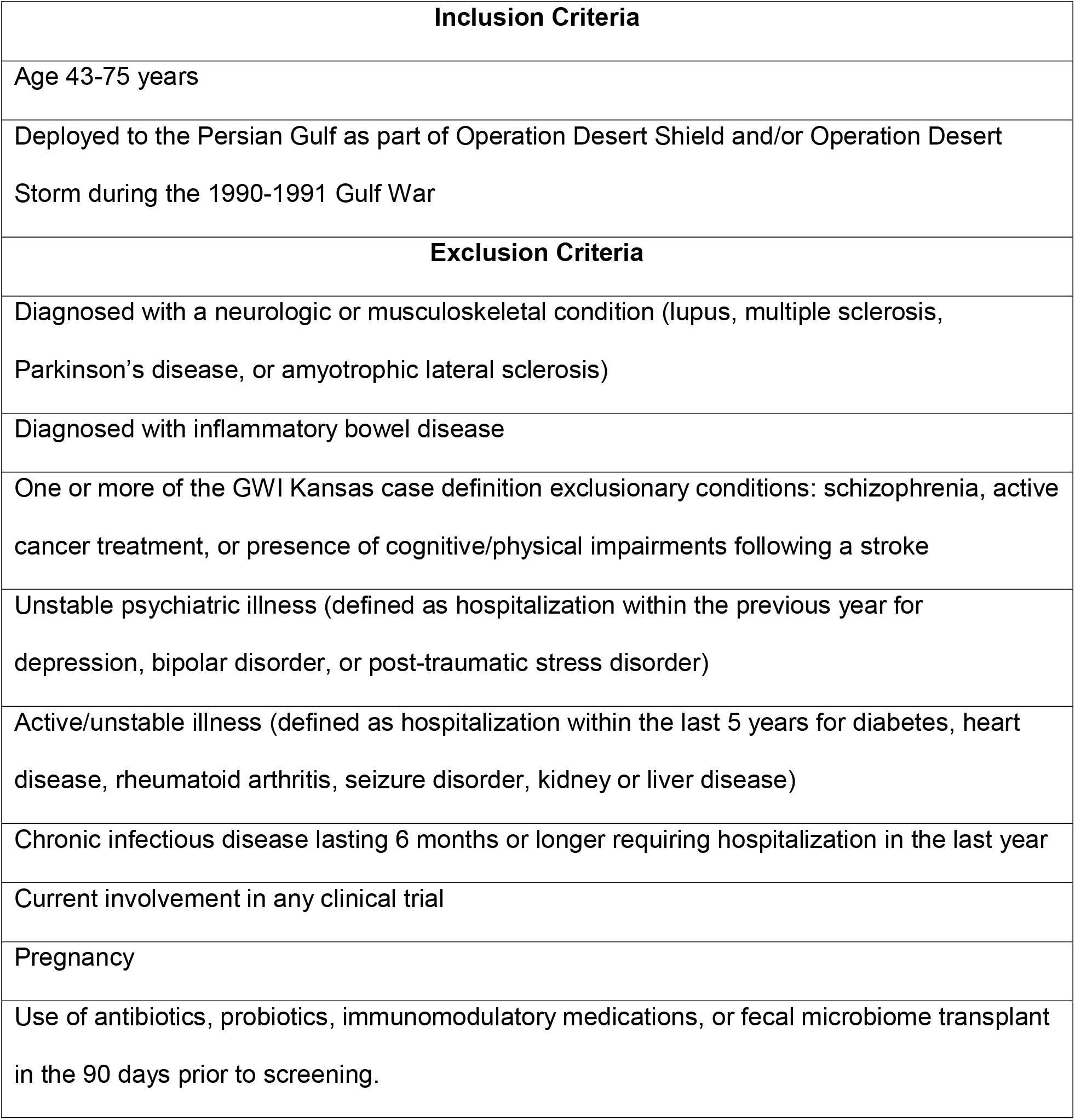
Study inclusion and exclusion criteria.

| **Inclusion Criteria** |
| --- |
| Age 43-75 years |
| Deployed to the Persian Gulf as part of Operation Desert Shield and/or Operation Desert Storm during the 1990-1991 Gulf War |
| **Exclusion Criteria** |
| Diagnosed with a neurologic or musculoskeletal condition (lupus, multiple sclerosis, Parkinson's disease, or amyotrophic lateral sclerosis) |
| Diagnosed with inflammatory bowel disease |
| One or more of the GWI Kansas case definition exclusionary conditions: schizophrenia, active cancer treatment, or presence of cognitive/physical impairments following a stroke |
| Unstable psychiatric illness (defined as hospitalization within the previous year for depression, bipolar disorder, or post-traumatic stress disorder) |
| Active/unstable illness (defined as hospitalization within the last 5 years for diabetes, heart disease, rheumatoid arthritis, seizure disorder, kidney or liver disease) |
| Chronic infectious disease lasting 6 months or longer requiring hospitalization in the last year |
| Current involvement in any clinical trial |
| Pregnancy |
| Use of antibiotics, probiotics, immunomodulatory medications, or fecal microbiome transplant in the 90 days prior to screening. |

### Enrollment

Veterans meeting eligibility criteria attended an in-person enrollment session. During the enrollment visit, participants completed questionnaires assessing their medical history and lifestyle including smoking history, medications, alcohol use, and comorbidities. Participants also completed the National Cancer Institute’s Dietary History Questionnaire III (DHQ III) (15,16) and a questionnaire assessing their known exposures to chemicals, biologics, and pharmaceuticals during their Gulf War deployment as well as their branch of service.

Lastly, participants completed the Kansas GWI case definitions assessment. Participants self-reported individual symptoms within the six GWI symptom domains (fatigue, pain, gastrointestinal, cognition, skin, and respiratory) and rated the severity of those symptoms as no, mild, moderate, or severe impacts on daily life in the previous six months. Deployed GWVs with GWI must have endorsed one or more moderate to severe symptoms in the fatigue domain and at least two of the other five symptom domains. Participants not meeting these criteria were classified as controls.

### Sample collection and study follow-up

Stool samples were collected weekly for eight weeks. Participants were provided with stool collection kits and were trained by the research team on how to properly collect, store, and ship specimens. Participants collected samples within a five-day window each week and were asked to ship specimens overnight within 24 hours of collection. Samples not received within 72 hours were rejected. The study team called participants weekly to remind them to collect and ship their samples. During the weekly check-in, participants were asked about antibiotic or probiotic use in the last week, about gastrointestinal symptoms, and if they had questions or comments for the study team.

### DNA extraction and sequencing

The full laboratory methods for this study have previously been published.(14,17) Briefly, total genomic DNA was extracted using a phenol:chloroform:isoamyl alcohol and bead-beating protocol with additional enzymatic lysis containing mutanolysin, lysostaphin, and lysozyme to assist in lysing gram-positive cell walls. Samples were purified using the NucleoSpin Gel and PCR cleanup kit according to the manufacturer’s directions (Macherey-Nagal, Germany) and stored at -80°C. DNA was quantified using the Qubit dsDNA kit (Invitrogen, Waltham, MA, USA) on the Biotek Synergy HTX (Biotek Instruments, Winooski, VT, USA). Samples were then sequenced using 16S rRNA sequencing of the V4 region on the Illumina MiSeq at the University of Wisconsin Biotechnology Center. Purified DNA was normalized to 5 ng/μ L, amplified using barcoded primers for the 16S V4 region, and sequenced using 2×250 paired end reads.

### Microbial analysis and statistics

Raw sequences were processed into amplicon sequencing variants (ASVs) using QIIME2(18) following the “Moving Pictures” protocol.(19) DADA2(20) was used for the quality control steps. Taxonomy was assigned using the GreenGenes(21) database and assigned to the genus level whenever possible. Statistical analyses were conducted using R version 4.1.0.(22) Alpha diversity was assessed using the Shannon, Inverse Simpson’s diversity indices; richness was assessed using the Chao1 index. The Bray-Curtis dissimilarity matrix, which was used to assess beta diversity, was visualized using non-metric multidimensional scaling (NMDS). Permutational Analysis of Variance (PERMANOVA) was used to estimate associations between GWI status and beta diversity.(23) In addition to GWI vs controls, alpha and beta diversity between those with and without gastrointestinal symptoms was also assessed. The Quasi-Conditional Association Test and Generalized Estimating Equation (QCAT-GEE) (24) was used to test for differentially abundant taxa (subset to the 100 most prevalent ASVs) between those with GWI, after adjustment for age, added sugars, the presence of gastrointestinal symptoms, pesticide exposure, and the Healthy Eating Index – 2015 (HEI). The HEI is a measure of overall diet quality (independent of food quantity) and is scored by assigning points to 13 dietary components with a maximum of 100 points.(25). A QCAT-GEE analysis was also done by HEI. The Benjamini-Hochberg correction for the false discovery rate (FDR) (26) was applied (α=0.05). QCAT-GEE provides three tests for differences by GWI status: the positive part (differences in abundance of each taxa), the zero part (differences in the presence/absence of taxa), and the two part (combining the positive and zero parts). A p-value of ≤0.05 was used for all statistical tests.

## Results

A total of 69 GWVs, 36 with GWI and 33 healthy controls, were included in the final analysis (Figure 1). The 69 patients provided 498 stool samples over eight weeks. The average age for the full cohort was 56.3 years; those with GWI were younger (54.2 years) than those without GWI (58.8 years, p=0.006). Six (9%) participants were female, three in each group. A majority of participants were members of the Army (N=46, 67%), followed by the Marines (N=12, 17%), Navy and Air Force (each N=5, 7%), and the Coast Guard (N=1, 1%). Fifty-nine (86%) participants identified as White. Full demographics data can be found in Table 2. Those with GWI reported significantly more moderate/severe symptoms across all symptom domains (Table 3). Twelve members of the GWI group reported experiencing three symptoms, 13 reported four symptoms, eight reported five symptoms, and three reported six symptoms. Six of those without GWI reported no symptoms, 11 reported one symptom, 12 reported two symptoms, two reported three symptoms, and two reported four symptoms. All exposures except vaccinations were more prevalent in the GWI group (Table 4).

**Figure 1.**
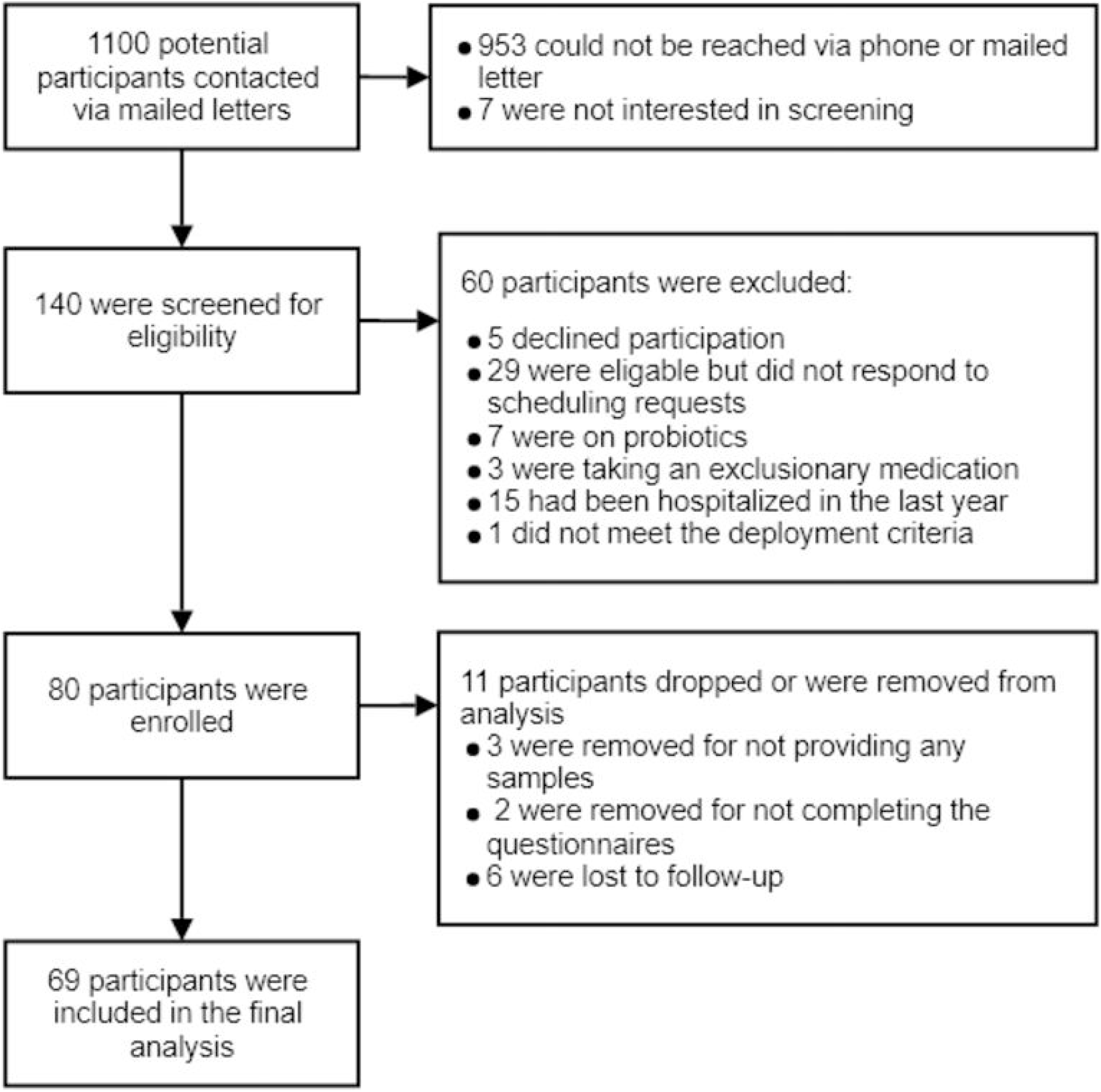
Flowchart of study enrollment.

**Table 2.** Participant demographics by age group.

|  | Full Cohort (N=69) | GWI (N=36) | Control (N=33) | p-value |
| --- | --- | --- | --- | --- |
| Age (years) | 56 | 59 | 54 | 0.006* |
| Gender |  |  |  |  |
| Male | 63 (91%) | 33 (92%) | 30 (91%) |  |
| Female | 6 (9%) | 3 (8%) | 3 (9%) | 0.91 |
| Branch |  |  |  |  |
| Army | 46 (67%) | 26 (72%) | 20 (67%) |  |
| Navy | 5 (7%) | 1 (3%) | 4 (13%) |  |
| Marines | 12 (17%) | 7 (19%) | 5 (17%) |  |
| Air Force | 5 (7%) | 2 (6%) | 3 (10%) |  |
| Coast Guard | 1 (1%) | 0 (0%) | 1 (3%) | 0.41 |
| Race* |  |  |  |  |
| White | 59 (86%) | 31 (86%) | 28 (85%) |  |
| Other | 8 (12%) | 4 (11%) | 4 (12%) |  |
| Preferred not to answer | 2 (3%) | 1 (3%) | 1 (3%) | 0.99 |

**Table 3.** Reported moderate/severe symptoms and GWI domain by group.

|  | Full Cohort (N=69) | GWI (N=36) | Control (N=33) |
| --- | --- | --- | --- |
| Symptom Domain |  |  |  |
| Fatigue | 46 (67%) | 36 (100%) | 10 (30%) |
| Pain | 36 (52%) | 33 (92%) | 3 (9%) |
| Neurologic | 43 (62%) | 31 (86%) | 12 (36%) |
| Skin | 12 (17%) | 10 (28%) | 2 (6%) |
| Respiratory | 14 (20%) | 11 (31%) | 3 (9%) |
| Gastrointestinal | 26 (38%) | 23 (64%) | 3 (9%) |
| Number of Symptoms |  |  |  |
| Zero | 6 (7%) | 0 (0%) | 6 (18%) |
| One | 11 (16%) | 0 (0%) | 11 (33%) |
| Two | 12 (17%) | 0 (0%) | 12 (36%) |
| Three | 14 (20%) | 12 (33%) | 2 (6%) |
| Four | 15 (22%) | 13 (36%) | 2 (6%) |
| Five | 8 (12%) | 8 (22%) | 0 (0%) |
| Six | 3 (4%) | 3 (8%) | 0 (0%) |

**Table 4.**
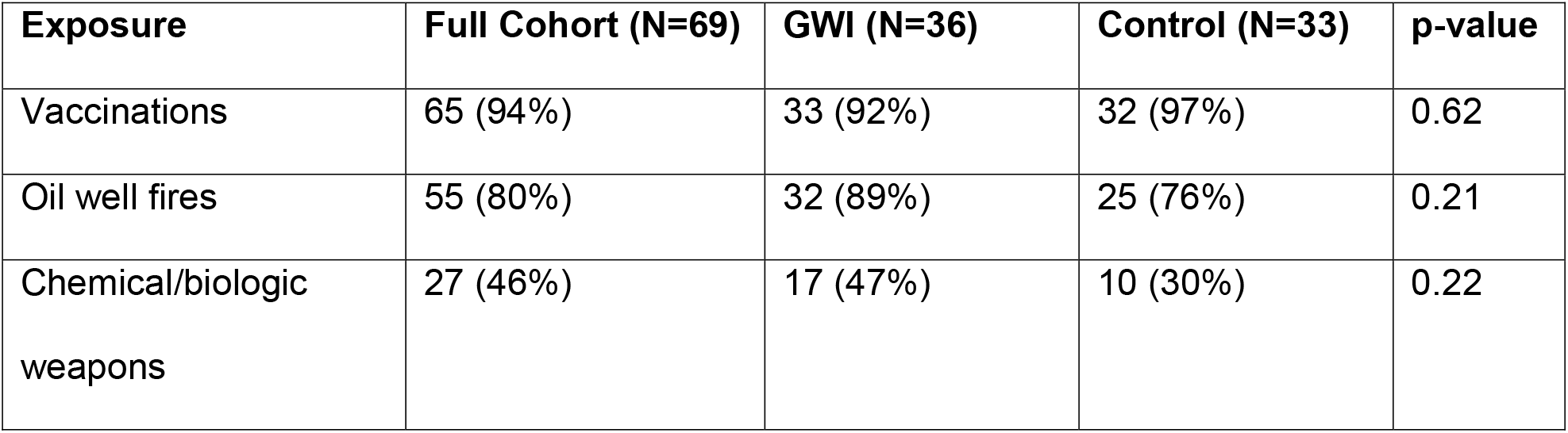
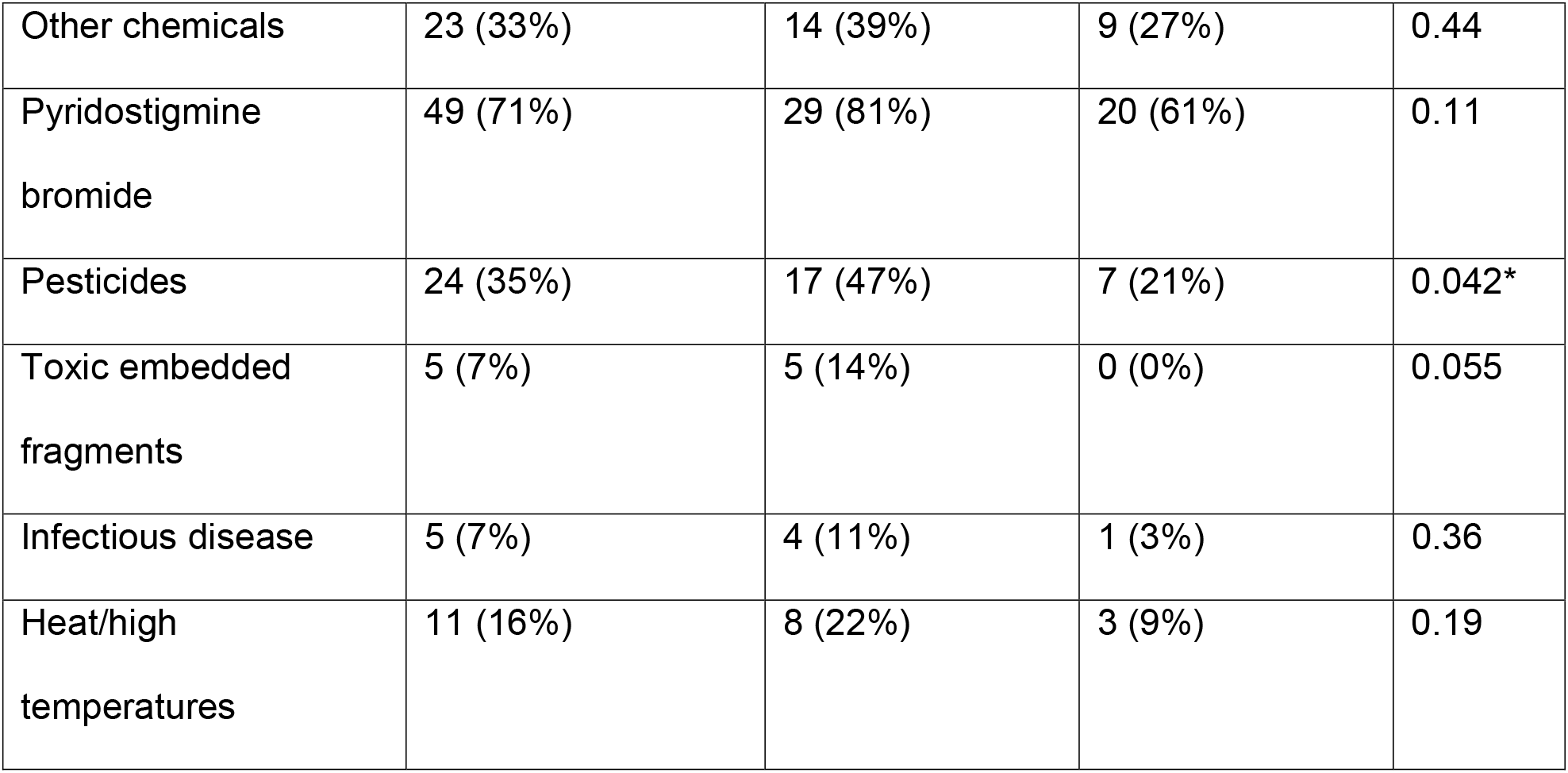
Reported exposures by group.

Overall, participants in both the control and GWI groups reported eating diets similar to the U.S. national average (59 pts) with the average Healthy Eating Index (HEI) at 61 pts (GWI: 58.2, Control: 63.8) using the National Cancer Institute’s Dietary History Questionnaire III (DHQ III). Those in the GWI group did have a poorer diet and had a borderline significantly lower score (p-value:0.0535). Those in the GWI group also reported eating diets containing significantly more added sugars (GWI: 17.2 tsp/day, Control: 11.0 tsp/day, p-value: 0.044) and well above the recommended amount of added sugar (Table 5). No other significant differences were observed between the groups. Additionally, the mean dietary intakes for both groups fell below the USDAs recommended daily intakes for all macronutrients.(27)

**Table 5.** Mean dietary data by group.

|  | Full Cohort<br>(N=69) | GWI (N=36) | Control<br>(N=33) | p-value | Recommended<br>Value* |
| --- | --- | --- | --- | --- | --- |
| Healthy Eating<br>Index | 61.0 | 58.2 | 63.8 | 0.054 | 100 is highest<br>score |
| Fiber |  |  |  |  |  |
| Total Dietary<br>Fiber (grams) | 18.3 | 18.4 | 18.2 | 0.95 | 31 g/day |
| Soluble Dietary<br>Fiber (grams) | 7.1 | 7.0 | 7.2 | 0.88 | No set<br>recommendation<br>s |
| Insoluble Dietary<br>Fiber (grams) | 11.2 | 11.3 | 11.0 | 0.86 | No set<br>recommendation<br>s |
| Total Fruit (cups) | 1.0 | 1.1 | 1.0 | 0.81 | 2.0 |
| Total Vegetables<br>(cups) | 1.5 | 1.7 | 1.4 | 0.27 | 3.0 |
| Total Legumes<br>(cups) | 0.1 | 0.1 | 0.1 | 0.74 | Approx. 0.3<br>cups/day or 2<br>cups/week |
| Whole grains<br>(ounces) | 0.8 | 0.8 | 0.8 | 0.91 | >3.5 |
| Refined grains<br>(ounces) | 3.4 | 3.9 | 3.0 | 0.13 | <3.5 |
| Total protein<br>foods (ounces) | 5.4 | 5.7 | 5.0 | 0.46 | 6.0 |
| Total Dairy<br>(cups) | 2.1 | 2.4 | 1.8 | 0.2 | 3.0 |
| Added sugars<br>(teaspoons) | 14.2 | 17.2 | 11.0 | 0.044 | <10% of<br>calories/day or |
|  |  |  |  |  | <13 tsp |
| Energy from total fat (%kcal) | 35.4% | 35.6% | 35.2% | 0.86 | 20-35% |
| Energy from carbohydrates (%kcal) | 44.8% | 44.9% | 44.6% | 0.9 | 45-65% |
| Energy from protein (% kcal) | 15.9% | 15.4% | 16.4% | 0.32 | 10-35% |
| Energy from saturated fats (% kcal) | 12.1% | 12.0% | 12.1% | 0.92 | <10% |

The gut microbiota for all participants were dominated by members of the Firmicutes and Bacteroides phyla. Participants in the control group who reported experiencing moderate/severe gut-related symptoms had a very different gut microbial composition compared to all other groups. The ratio of Bacteroidetes to Firmicutes (B/F ratio) for the full cohort was 54.9% to 38.0% (Figure 2). Actinobacteria (2.3%), Verrucomicrobia (2.5%), and Proteobacteria (1.8%) were the only other phyla consisting of over 1% of the ASVs.

**Figure 2.**
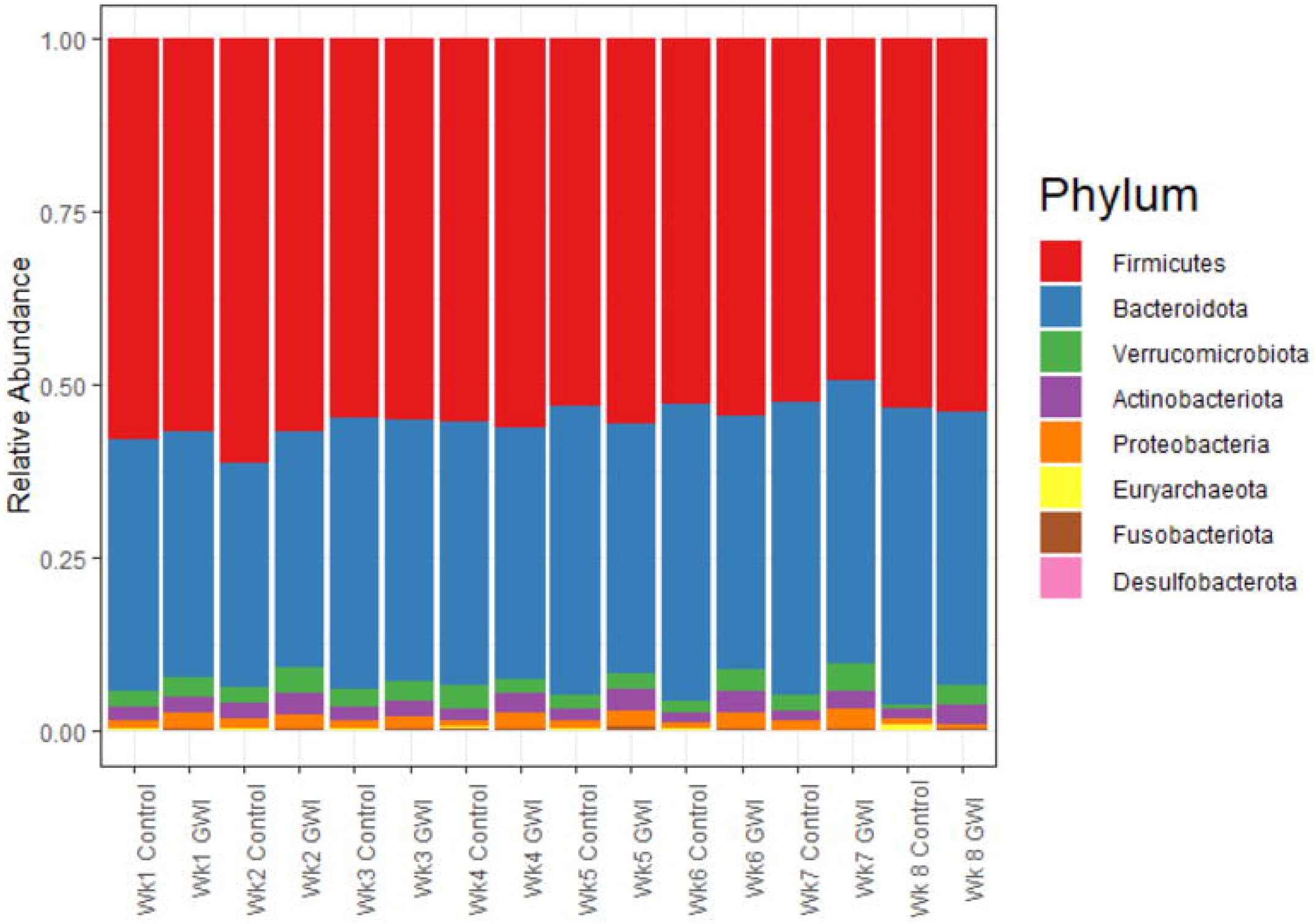
Relative abundance bar plot of the top 8 phyla by aggregated by week and GWI status.

The Shannon, Inverse Simpson, and Chao1 alpha diversity index measures all showed a very stable microbiome over time, regardless of GWI status. Linear mixed-effects modeling of the three alpha diversity measures confirmed these findings (p=0.28) (Figure 3). While no differences between groups were statistically significant, a visual inspection of the data shows slightly lower alpha diversity amongst those reporting moderate to severe gut symptoms regardless of GWI status (Figure 4).

**Figure 3.**
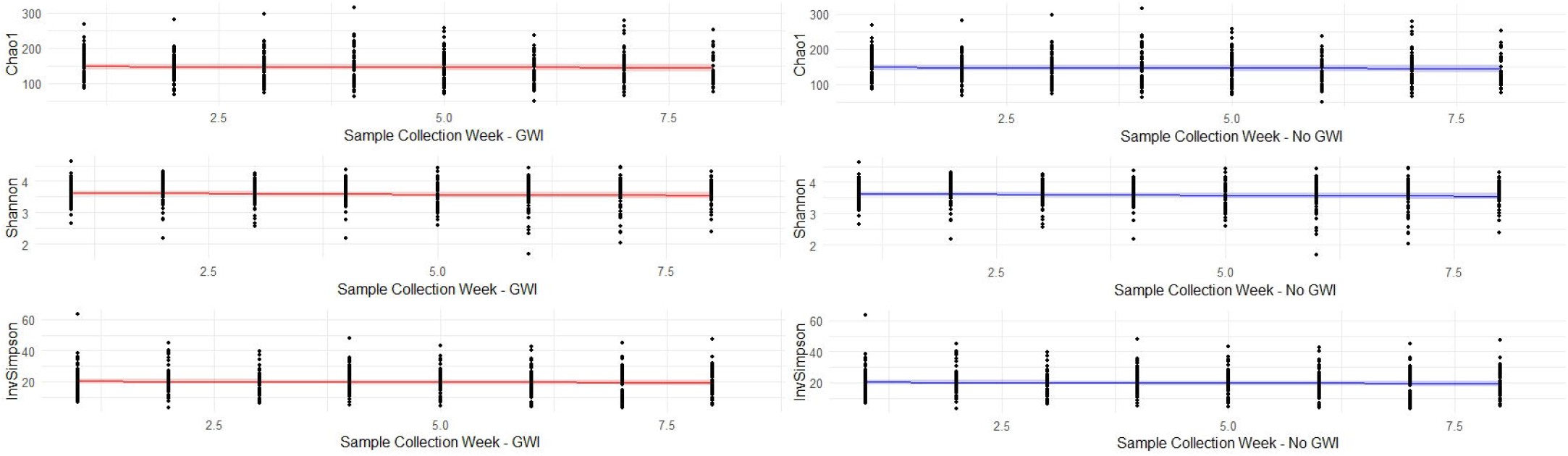
Linear mixed-effects modeling of alpha diversity by week. Red line is those with GWI and blue line is the controls. Lighter bands represent the 95% confidence interval of the regression line.

**Figure 4.**
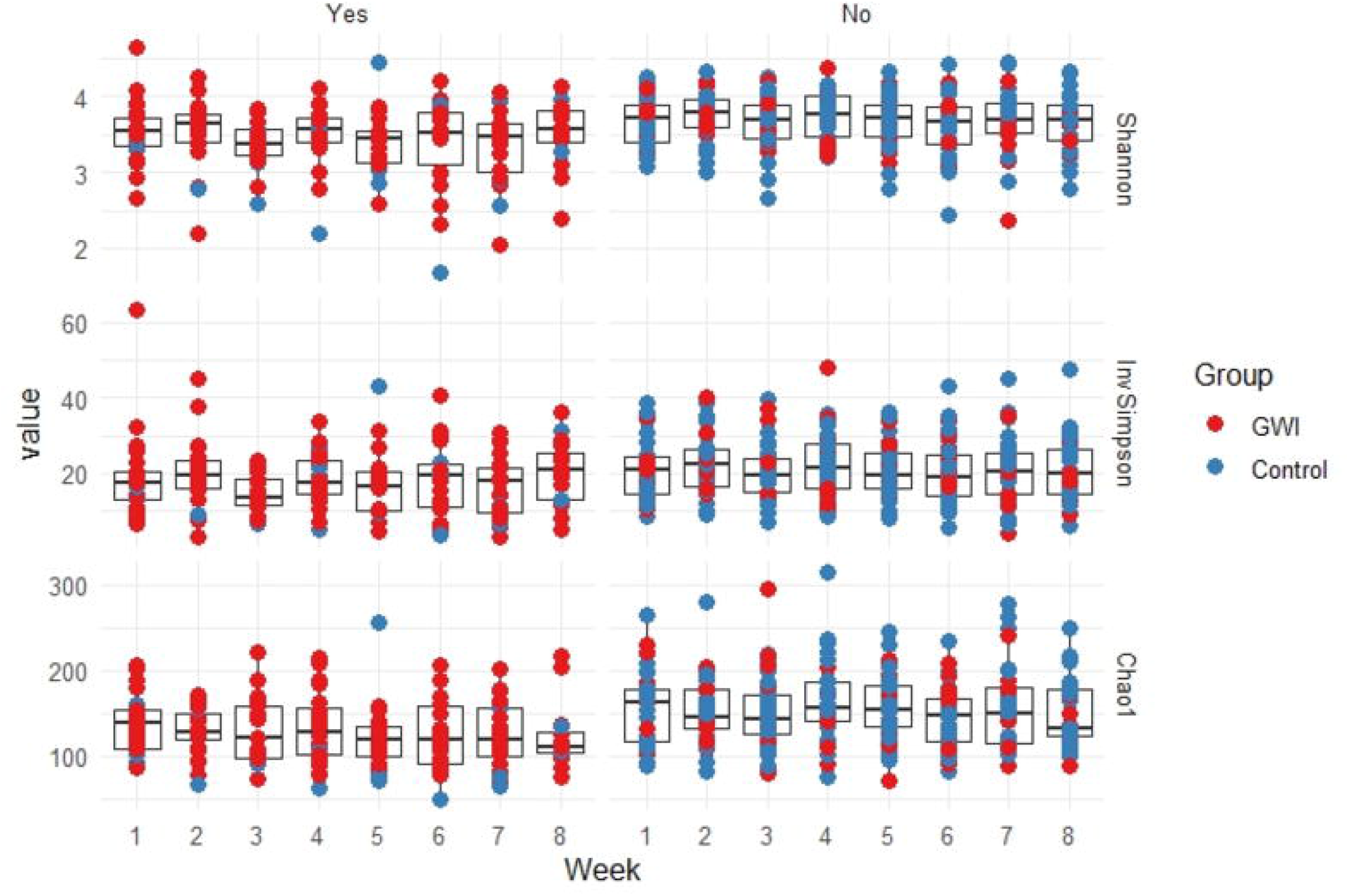
Alpha diversity by week and GWI status stratified by the presence of moderate/severe gastrointestinal symptoms (Yes/No).

To assess beta diversity, the Bray-Curtis dissimilarity was calculated and visualized using NMDS (Figure 5). Visual inspection showed a great deal of overlap of the ellipses indicating the centroids are in similar places; however, the PERMANOVA analysis indicated group status (GWI vs. controls, and GWI +GI, GWI, Control +GI, and control) significantly explained 1.2% and 3% of the variation in the data respectively (p-value:0.001 for both). However, PERMANOVA assumes homoscedasticity and the data showed significantly different dispersions (p=0.002 via betadisper in vegan). To confirm results, analysis of similarities (ANOSIM) was done which also found GWI status to significantly impact the gut microbiota as well (p <0.0001). The microbiota was very stable over the duration of the study with “week” not being associated with beta diversity using either PERMANOVA or ANOSIM (p=1.0 and 0.999 respectively).

**Figure 5.**
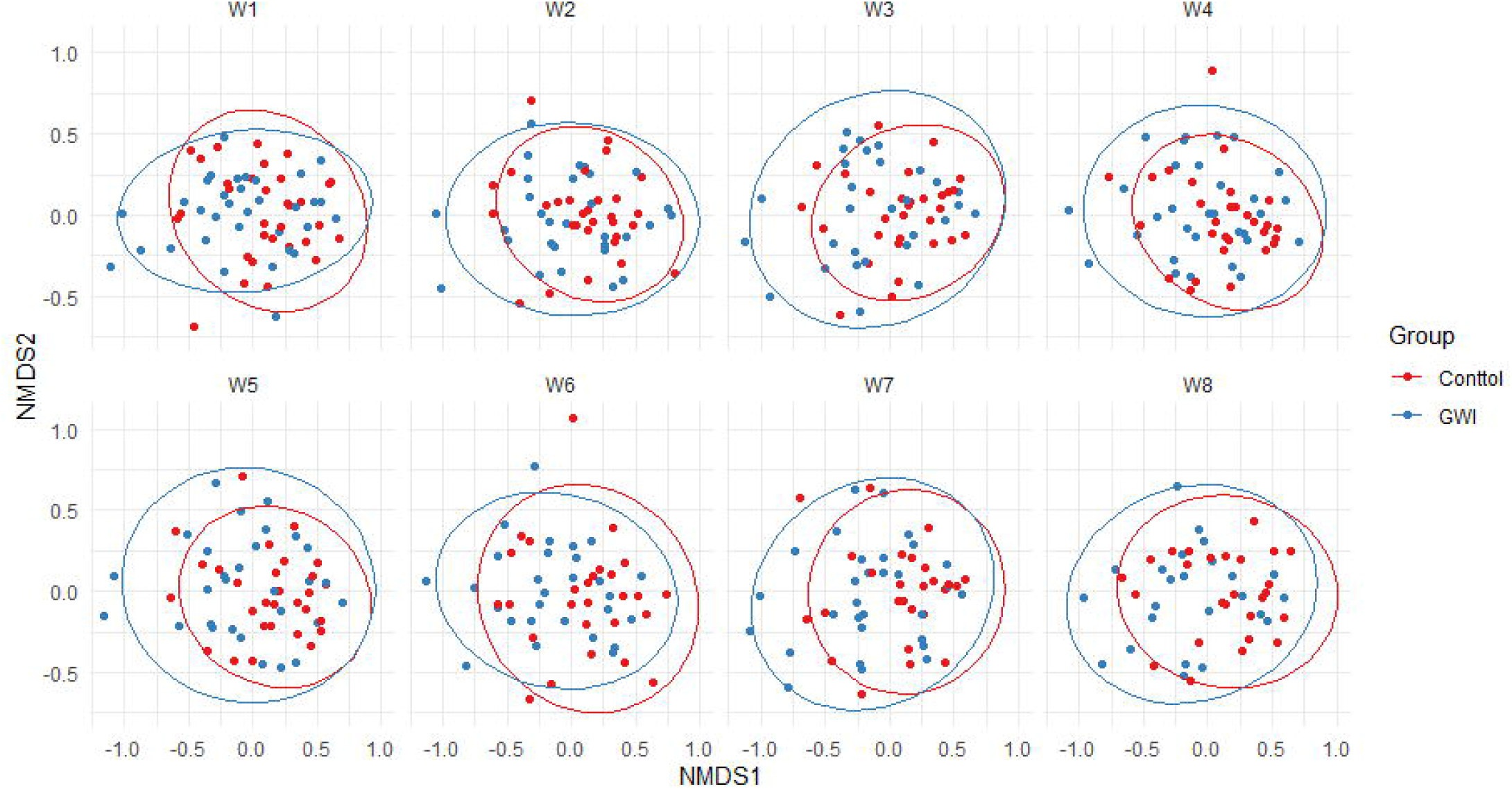
NMDS plot of the Bray-Curtis dissimilarity matrix by week and GWI status.

The QCAT-GEE test for differential lineages identified 9 lineages at the family level that were significantly different between those with GWI and the healthy controls. This model was adjusted for the subjects’ age, the presence of gastrointestinal symptoms, pesticide exposure during deployment, HEI score, and added sugar consumption (in teaspoons). Three lineages (Lachnospiraceae, Oscillospiraceae, and Ruminococcaceae) were more prevalent in those without GWI and were significant across the positive-, zero-, and two-part tests (Table 6). Additionally, we tested for whether there were differences in lineages associated with eating habits (HEI score) those experiencing moderate to severe gastrointestinal symptoms (adjusted for GWI status) (Table 7). Lineages associated with the Eggerthellaceae, Erysipelatoclostridiaceae, Lachnospiraceae, and Oscillospiraceae families were all significant across the positive-, zero-, and two-part tests with Erysipelatoclostridiaceae and Lachnospiraceae associated with decreasing HEI scores.

**Table 6.** Bacterial taxa associated with GWI adjusted for covariates at the family level.

| Taxa | Positive Part | Zero Part | Two-Part | Direction* |
| --- | --- | --- | --- | --- |
| Acidaminococcaceae | 0.001 | 0.849 | 0.008 | ↑ |
| Eggerthellaceae | 0.446 | 0.028 | 0.059 | ↑ |
| Erysipelotrichaceae | 0.0735 | 0.002 | 0.012 | ↓ |
| Lachnospiraceae | 0.001 | 0.0001 | 0.001 | ↓ |
| Marinifilaceae | 0.0001 | 0.214 | 0.0001 | ↑ |
| Oscillospiraceae | 0.007 | 0.004 | 0.0001 | ↓ |
| Peptostreptococcaceae | 0.986 | 0.015 | 0.035 | ↓ |
| Ruminococcaceae | 0.002 | 0.041 | 0.001 | ↓ |
| Veillonellaceae | 0.019 | 0.317 | 0.061 | ↑ |

*P*-values reported have FDR correction applied.
\*Up arrows indicate higher abundance in those with GWI, down arrows indicate higher abundance in controls

**Table 7.**
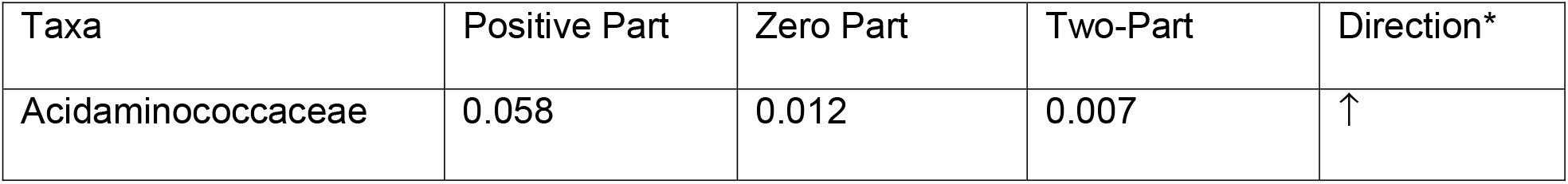
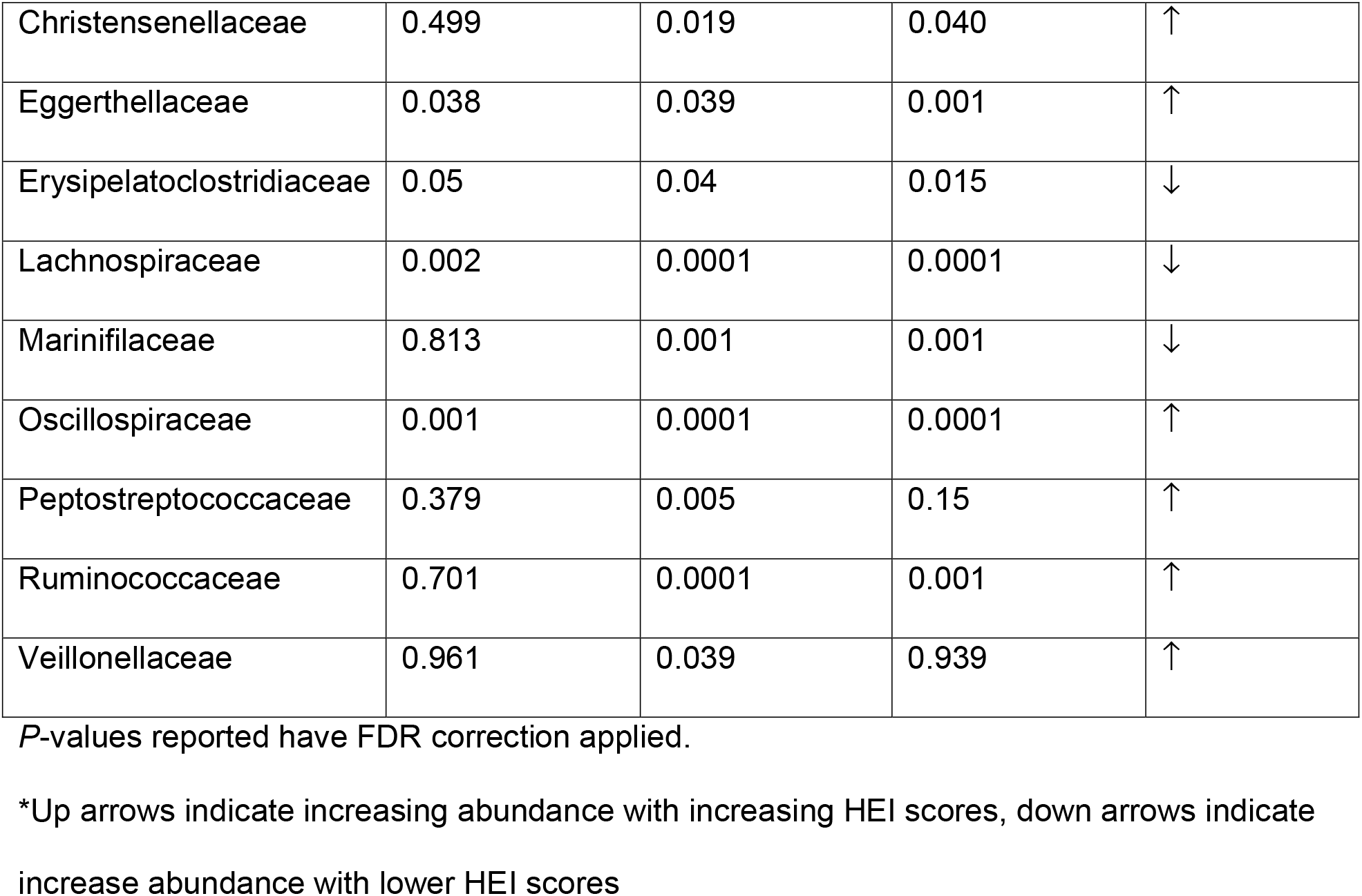
Bacterial taxa associated with HEI scores among those with gastrointestinal symptoms adjusted for GWI status at the family level.

| Taxa | Positive Part | Zero Part | Two-Part | Direction* |
| --- | --- | --- | --- | --- |
| Acidaminococcaceae | 0.058 | 0.012 | 0.007 | ↑ |
| Christensenellaceae | 0.499 | 0.019 | 0.040 | ↑ |
| Eggerthellaceae | 0.038 | 0.039 | 0.001 | ↑ |
| Erysipelatoclostridiaceae | 0.05 | 0.04 | 0.015 | ↓ |
| Lachnospiraceae | 0.002 | 0.0001 | 0.0001 | ↓ |
| Marinifilaceae | 0.813 | 0.001 | 0.001 | ↓ |
| Oscillospiraceae | 0.001 | 0.0001 | 0.0001 | ↑ |
| Peptostreptococcaceae | 0.379 | 0.005 | 0.15 | ↑ |
| Ruminococcaceae | 0.701 | 0.0001 | 0.001 | ↑ |
| Veillonellaceae | 0.961 | 0.039 | 0.939 | ↑ |

*P*-values reported have FDR correction applied.
\*Up arrows indicate increasing abundance with increasing HEI scores, down arrows indicate increase abundance with lower HEI scores

## Discussion

This longitudinal cohort study of GWVs with and without GWI found participants with GWI had significantly different microbiomes from those without GWI. Furthermore, the gut microbiota was relatively stable over the eight weeks, regardless of GWI status, which was not surprising given no intervention was part of the study. The gut microbiota of both those with and without GWI consisted almost entirely of members of the Firmicutes and Bacteroidetes phyla across all time points. While there was no difference in alpha diversity between the groups, there was a difference in beta diversity by both group and the presence of gut symptoms. Similar to alpha diversity, beta diversity was not impacted by sampling week. We also identified several lineages that differed significantly by GWI status including members of the families Lachnospiraceae, Oscillospiraceae, and Ruminococcaceae which were all more prevalent in those without GWI. Last, it appears those participants reporting gastrointestinal symptoms with healthier eating habits had differing gut microbiota from those with less healthy diets.

While limited data on the impact of GWI and the gut microbiome exists, several studies have observed greater abundances in Lachnospiraceae in those without GWI.(13) However, in a pilot study of Veterans with GWI, Ruminococcaceae was found to be associated with GWI, while in our study, it was associated with the healthy participant group. One potential reason for this difference is the smaller sample sizes observed in the pilot study of Veterans with GWI (n=16 with stool samples, 3-5 per group).(13) Other studies assessing the gut microbiome in GWI mouse models have also found Ruminococcaceae to be more abundant in the healthy controls.(12),(28) Previous research on Crohn’s disease and inflammatory bowel disease (IBD) have found members of the Ruminococcaceae family to be decreased in those with the disease state,(29) consistent with our findings. Several members of the Ruminococcaceae family are important in microbe-mediated carbohydrate metabolism breaking down starches and cellulose in the lower GI tract.(30) They are critical producers of acetate(31), a necessary short-chain fatty acid (SCFA) for gut health as well as serving as a critical nutrient for butyrate producing bacteria in the gut.(32),(33) While not assessed in our study, acetate is a potential mediator of the gut-brain axis.(34) Animals models of multiple sclerosis (MS) have shown acetate supplementation can help reduce clinical symptoms associated with MS in mice.(35) In our cohort, 31 (86.1%) of those with GWI endorsed experiencing moderate to severe neurologic symptoms. Lachnospiraceae has also been shown to be in decreased abundance in those with GWI in both human and mouse models.(13),(28),(36) Additionally, it has been shown to be reduced in those with ulcerative colitis(37), ME/CFS(38), and Crohn’s disease(39). Members of the Lachnospiraceae family are also key carbohydrate metabolizers producing butyrate.(37)

SCFAs, including butyrate and acetate, are crucial to gut health. They are the end products of carbohydrate metabolism primarily produced through anaerobic fermentation of dietary fiber in the gut. They also promote epithelial barrier function, cell proliferation, and act as a nutrient source for many beneficial microbes in the gut.(32) Dietary interventions, either through food or supplements, to increase SCFAs have been proposed as potential treatments for a wide range of gut disorders including GWI. Prior studies have shown Western and high fat diets are associated with GWI.(12),(28),(40) Additionally, high sugar diets have been associated with decreased SCFAs, increased gut membrane permeability, and decreased microbial diversity in mouse models of ulcerative colitis.(41) In our study, the diets of all participants fell below the daily recommended amounts of dietary fiber, vegetables, whole grains, and fruits. Additionally, participants in both groups met or exceeded the recommended amount of energy from fats (including saturated fats). Those with GWI consumed significantly more added sugars than those without GWI and had a slightly lower HEI score. Poor overall diet was associated with ten differentially abundant lineages, most notable Lachnospiraceae and Ruminococcaceae. Ruminococcaceae increased with higher HEI scores, while Lachnospiraceae trended towards greater abundance in those with lower scores. However, 33 ASVs were associated with this lineage, with some being more abundant in those with higher HEI scores (*Dorea, Anaerostipes, Lachnoclostridium*, and *Ruminococcus)* and some more abundant in those with lower scores (*Agathobacter, Blautia*, and *Rosburia*). Prior research has shown Lachnospiraceae to have a complicated relationship to gut health with diet having a large influence on abundance.(37) This, in conjunction with the low levels of Lachnospiraceae and Ruminococcaceae found in those with GWI, indicates dietary interventions focused on improving diet and increasing SCFA production may benefit Veterans with GWI.

Our study has several strengths and limitations. This is the first longitudinal cohort study on GWI assessing the role of the gut microbiome and diet. We had a low dropout rate with 86% of participants completing the study and a majority completing the dietary questionnaire (DHQ III). One limitation to the study was the DHQ III. While food frequency questionnaires like the DHQ III are standard means of collecting dietary data(42), it is cumbersome to complete and relies heavily on self-reported data of foods consumed over the last month. Studies have shown self-report dietary intakes underestimate the true energy consumption and individuals tend to report intakes closer to their perceived norms as opposed to actual consumption.(43) Participants in our cohort likely ate worse diets than reported.

## Conclusions

Future studies assessing the role of diet and the gut microbiome should strive to collect more detailed dietary data through the use of food diaries to reduce recall bias. Additionally, research building upon these findings would benefit from the use of shotgun metagenomics sequencing and the addition of metabolomics. Both metabolomics and metagenomic data would provide insight into the functional role these microbiota play in the gut of those with GWI. Furthermore, these methods would allow researchers to examine the role of SCFAs. Last, these findings along with the existing literature on GWI, point to the role of dietary interventions in alleviating the gastrointestinal symptoms frequently experienced by those with GWI. Studies assessing dietary and other interventions to manipulate the gut microbiome are needed to examine the safety and efficacy of treatments aimed at reducing GWI symptoms.

## Data Availability

Raw sequence reads and corresponding metadata for all samples described in the paper have been deposited in the NCBI Sequence Read Archive (SRA) under accession no. PRJNA798786. Further information requests should be directed to the corresponding author.

## Acknowledgments

The authors would like to thank Catherine Shaughnessy for her help during the recruitment, enrollment, and data collection phases of this study.

